# Synthetic data for privacy-preserving clinical risk prediction

**DOI:** 10.1101/2023.05.18.23290114

**Authors:** Zhaozhi Qian, Thomas Callender, Bogdan Cebere, Sam M Janes, Neal Navani, Mihaela van der Schaar

## Abstract

Synthetic data promise privacy-preserving data sharing for healthcare research and development. Compared with other privacy-enhancing approaches - such as federated learning - analyses performed on synthetic data can be applied downstream without modification, such that synthetic data can act in place of real data for a wide range of use cases. However, the role that synthetic data might play in all aspects of clinical model development remains unknown. In this work, we used state-of-the-art generators explicitly designed for privacy preservation to create a synthetic version of the UK Biobank before building prognostic models for lung cancer under several data release assumptions. We demonstrate that synthetic data can be effectively used throughout the modelling pipeline even without eventual access to the real data. Furthermore, we show the implications of different data release approaches on how synthetic data could be deployed within the healthcare system.

## Introduction

Medical advances are predicated on the availability of high-quality data, leading to an increasing emphasis on data sharing in both industry and academia. Nevertheless, the sensitivity of medical data is such that it is usually tightly controlled and subject to country-specific legal constraints^1,2^. Consequently, data access remains complex, inconsistent, costly, and time-consuming^3–5^. Synthetic data have been recognized as a promising solution, coupling privacy-preservation with sufficient quality for analysis^6,7^. Generated by algorithm, synthetic data can maintain the statistical properties and distributions of an original dataset but represent newly created participants.

Compared with other privacy enhancing technologies, such as federated learning^8,9^, synthetic data has an unique advantage: all downstream analytical and ML algorithms can be applied to synthetic data in the same way they are applied to real data. The seamless switch between real and synthetic data allows the data user to apply statistical and ML algorithms without replacing or overhauling these tools. Hence, the use cases of synthetic data span the whole life cycle of a data science project, from exploratory data analysis (EDA)^10^ and model development - including dimensionality reduction, cluster analysis, hypothesis testing, feature selection, hyperparameter tuning - through model selection and training.

Two approaches are commonly proposed for deploying synthetic data: “*no-release*” and “*delayed-release*”. Under a *no-release* approach, the data controller only ever releases synthetic data to the user. This allows a variety of applications such as running data science competitions^11^ or the evaluation of new software prior to deployment. Under a *delayed-release* paradigm, the data controller initially makes synthetic data available to a user, followed by the delayed release of the real data. A *delayed-release* approach supports multiple use cases. Users could accelerate and de-risk analytical projects as the approval process for real data access is often lengthy, such that many analyses can take months or years to start. Further, the quality and usefulness of any dataset, particularly real-world electronic health records, is often unclear in advance. By using a synthetic version initially, a data user can better understand whether the real data can support the proposed analyses, and identify where there may be issues with the real data that require addressing.

Both of these deployment paradigms require synthetic data that mirrors the conditional distribution between features and outcomes of interest, as well as the relationships between different features. Consequently, any method to generate synthetic data should achieve two goals: imitating the statistical and joint distributions in the real data, and ensuring that the privacy of those present in the original data is preserved. However, these two goals are sometimes in conflict with each other, leading to a trade-off between the usefulness of synthetic data and its privacy^12,13^. At its extreme, a synthetic data generator could memorise an individual’s features and return these in a synthetic dataset^14,15^. Standard generative models are focussed on the first goal of ensuring distributional similarity, while neglecting the second^16,17^. As a remedy, several approaches explicitly designed to allow control over an explicit privacy guarantee have been developed^18–21^. Most commonly, this involves the introduction of noise during the training of a synthetic data generator, such that the generator is presented with a blurred version of reality. However, the additional noise will inevitably perturb the true data distribution, reducing the usefulness of the synthetic data for certain analytical tasks.

Synthetic data have been shown capable of capturing the high-level marginal distributions and pairwise correlations between features^22–26^, as well as in training predictive models^27–30^. However, none of these studies have used synthetic data generators which explicitly control for privacy, a prerequisite in medicine, whilst whether and how synthetic data can be useful in other stages of the data science pipeline is still unexplored. Furthermore, existing studies often use small datasets and idealized prediction tasks for evaluation, raising questions about whether the results extrapolate to more complex and realistic settings^31,32^. In this study, we aimed to comprehensively examine the utility of synthetic data generated by state-of-the-art privacy-preserving generators at all stages of the clinical risk prediction pipeline. We show that existing synthetic data generation methods are of sufficient quality to support a broad range of uses under different access paradigms, empowering data controllers to deploy synthetic data for health research and development.

## Methods

### Data and study population

We used data from the UK Biobank, a large prospective cohort of half a million men and women recruited between 2006-10 from across the UK with ongoing follow-up^33^. Lung cancer screening is currently only considered in ever-smokers. Consequently, we included all 216,714 individuals in the UK Biobank without a previous diagnosis of lung cancer at baseline who self-reported as current or former smokers. Diagnoses of lung cancer during follow-up were determined through linked national cancer registry data^33^, right censored at 31st July, 2019.

### Variable selection and data pre-processing

We selected 26 candidate variables (Appendix Table 1) either causally linked to lung cancer or used in existing lung cancer prognostic models^34^. To manage missing data, we used multiple imputation with chained equations and predictive mean matching. Prior to analysis, as our synthetic data generators leverage neural networks, we normalised continuous variables such their values lay between 0 and 1. Categorical variables were one-hot encoded.

### Synthetic data generation

We used three synthetic data generators: DPGAN^19^, PATEGAN^20^, and ADSGAN^21^, all of which are specifically designed for privacy-preservation so are suitable for controlled healthcare datasets. We also considered PrivBayes^18^, but it did not scale to the size of the dataset. DPGAN, PATEGAN, and ADSGAN are based on generative adversarial networks^35^. This framework involves two opposing models: a generator that creates synthetic participants and a discriminator that attempts to predict whether these synthetic participants were part of the original dataset. Training continues until the data distributions learnt by the generator are indistinguishable from the original dataset.

For privacy preservation, DPGAN and PATEGAN implement algorithms for differential privacy^36^, whilst ADSGAN is specially designed to protect against re-identification attacks^21^. Differential privacy is a formal, mathematically-definable, notion based on the concept that participation in any database renders a risk of identification, such that it is the relative increase in risk of identification that is of importance^36^. By contrast, ADSGAN is specially designed to protect against re-identification (linkage) attacks - where publicly available data are combined to re-identify an individual - a type of privacy attack specifically highlighted in the European Union’s General Data Protection Regulation (GDPR)^1^.

To train the generators, we split our UK Biobank cohort 80:20 into a training 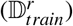 and test 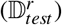 set. As this is a stochastic process, we repeated this ten times using different random seeds. We then generated 10 synthetic datasets, one from each trained generator, and aggregated them into one final synthetic dataset, 𝔻^*s*^ - a deep generative ensemble^37^. Given randomness in both generators, and the synthetic data produced by each generator, deep generative ensembling has been shown to improve the quality of the final synthetic dataset used^37^. This led to three main synthetic datasets, one each for DPGAN, PATEGAN, and ADSGAN. We set a privacy budget of *ε* = 1.0; remaining hyperparameters are available in the Appendix.

### Evaluating synthetic data for exploratory data analysis

We considered the performance of synthetic data for both descriptive analyses and dimensionality reduction. Descriptive analyses were comparative, showing the distributions of both continuous and categorical variables in the synthetic and real datasets. We used kernel density estimation with a Gaussian kernel to produce smoothed plots showing the distribution of continuous variables. For dimensionality reduction, we applied two widely-used techniques: principal component analysis (PCA)^38^ and K-means clustering^39^.

We performed PCA separately on the real training 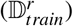 and synthetic (𝔻^*s*^) datasets, qualitatively comparing the profile of explained variance^40^. The profile of explained variance is an important tool to help decide the number of principal components. Ideally, the PCA model trained on the synthetic 𝔻^*s*^ should be close to a PCA model trained on the real dataset, 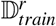, with a similar variance profile. For a quantitative comparison, we also evaluated the two trained PCA models on the real test set, 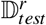, to measure the difference in their abilities to explain unseen real data in terms of the log-likelihood^41^. We repeated the analysis above for all synthetic data generators.

For K-means clustering, we performed the analysis separately on 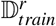 and 𝔻^*s*^ with clusters *k* = 2,…, 28. We then qualitatively compared the Bayesian Information Criterion (BIC) curves^42^ obtained from real and synthetic data to evaluate whether synthetic data can help the data user to select the optimal number of clusters. Finally, we applied the trained K-means algorithms to cluster the real test data 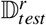. We evaluated the agreement in cluster assignment with the adjusted Rand index (ARI)^43^ and adjusted mutual information (AMI)^44^.

### Evaluating synthetic data for model development

We considered two central tasks in model development: feature selection and hyperparameter tuning. For feature selection, we first developed a Cox regression model on the real training data, 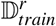, to obtain a “ground-truth” p-value, 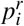for each candidate prognostic variable *X*_*i*_, that describes the strength of the association between the variable and lung cancer occurrence. We subsequently repeated this process on each synthetic dataset, 𝔻^*s*^, obtaining p-values 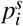. Keeping those prognostic variables that met a threshold for significance of *α* = 0.05, we created lists of selected features from the real and synthetic datasets. By comparing the variables that met this threshold - variables selected in both 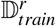and 𝔻^*s*^ were considered true positives, whilst those selected only in a synthetic dataset were considered false positives - we calculated the precision, recall, and the area under the receiver operating curve (AUROC) of feature selection using hypothesis testing with synthetic data.

For hyperparameter tuning, we trained a deep survival analysis model, DeepHit^45^. We used a randomised search approach for hyperparameter selection, generating a search grid containing 20 different settings. We split our synthetic data, 𝔻^*s*^, 80:20 into training and validation sets to train and evaluate DeepHit models with different hyperparameters before selecting the best configuration. Finally, we re-trained a model with the selected hyperparameters using the real dataset, 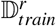 - imitating the delayed-release mode - and evaluated its performance on the real test dataset, 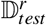, with the concordance index (C-index)^46^. As a baseline, we considered the average performance of the 20 settings on 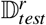.

### Evaluating synthetic data for model training

To explore the usefulness of synthetic data for model training, we used the train-on-synthetic, test-on-real approach in which we fitted a Cox regression model on the synthetic data, 𝔻^*s*^, with all candidate prognostic features and then evaluate its performance on the real, test, dataset 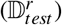. Using the 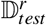 avoids potential data leakage issues that might occur with an evaluation on the real training datasets, 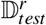. We considered model discrimination using the concordance index (C-index), as well as model performance and calibration using the Brier score^48^.

### Code and data availability

The code used in this project are available at https://github.com/vanderschaarlab/synthcity. UK Biobank data were used on license and cannot be directly shared; researchers can apply to the UK Biobank for access.

## Results

### Exploratory data analysis with synthetic data

#### Descriptive statistics

The descriptive characteristics of the synthetic and real datasets are shown in Table 1. Synthetic datasets generated with ADSGAN and PATEGAN both faithfully represented the training cohort. This extended to the complex multi-modal distribution shown in the number of cigarettes smoked per day, where individuals frequently reported values to the nearest five cigarettes (Figure 1). By contrast, DPGAN struggled to match the distributional characteristics of features, with notable inconsistencies amongst categorical variables, such as an individual’s ethnicity, personal history of cancer, COPD, or pneumonia, along with mode invention and mode collapse amongst key continuous variables.

**Table 1.** Descriptive characteristics of the original and synthetic data

|  | ADSGAN | PATEGAN | DPGAN | UK Biobank |
| --- | --- | --- | --- | --- |
| Age (mean, SD) | 57.41 (8.15) | 57.43 (8.34) | 54.81 (13.38) | 57.39 (7.93) |
| Sex - Female (n, %) | 82,693 (47.70) | 86,207 (49.72) | 84,264 (48.6) | 83,003 (47.88) |
| Ethnicity - White (n, %) | 164,398 (94.82) | 165,526 (95.48) | 5 (0.0) | 166,558 (96.45) |
| Highest qualification (n, %) |  |  |  |  |
| Degree | 48,413 (27.92) | 48,038 (27.71) | 46,302 (26.71) | 47,642 (28.00) |
| Some college | 14,045 (8.10) | 12,596 (7.27) | 38,521 (22.22) | 13,244 (7.78) |
| Post-secondary school | 29,134 (16.8) | 27,448 (15.83) | 31,442 (18.14) | 26,887 (15.80) |
| Secondary school | 46,463 (26.8) | 46,228 (26.66) | 2,312 (1.33) | 46,128 (27.11) |
| None of the above | 35,316 (20.37) | 39,061 (22.53) | 54,794 (31.61) | 36,249 (21.30) |
| Body mass index (mean, SD) | 27.54 (4.57) | 27.60 (5.19) | 35.86 (21.12) | 27.76 (4.78) |
| Smoking status (n, %) |  |  |  |  |
| Previous | 129,883 (74.92) | 132,522 (76.44) | 49,676 (28.65) | 131,822 (76.03) |
| Current | 43,488 (25.08) | 40,849 (23.56) | 123,695 (71.35) | 41,549 (23.97) |
| Age started smoking (mean, SD) | 17.31 (4.34) | 16.96 (4.69) | 18.39 (19.70) | 17.42 (4.33) |
| Years smoked (mean, SD) | 25.64 (12.85) | 25.39 (13.51) | 26.41 (19.34) | 26.32 (12.91) |
| Cigarettes per day (mean, SD) | 14.08 (10.54) | 15.04 (12.8) | 39.04 (42.59) | 18.20 (10.20) |
| Pack-years (mean, SD) | 18.96 (19.52) | 21.55 (23.43) | 73.86 (76.20) | 23.86 (18.90) |
| Personal history of cancer (n, %) | 19,080 (11.01) | 17,653 (10.18) | 173,368 (99.99) | 15,511 (8.95) |
| COPD (n, %) | 7,133 (4.11) | 4,872 (2.81) | 173,366 (99.99) | 5,310 (3.07) |
| Family history of lung cancer (n, %) | 31,495 (18.17) | 22,072 (12.73) | 173,371 (100.0) | 23,144 (13.6) |
| Pneumonia (n, %) | 4,079 (2.35) | 3,201 (1.85) | 173,367 (99.99) | 2,653 (1.53) |
| Asthma (n, %) | 24,780 (14.29) | 21,646 (12.49) | 172,996 (99.78) | 20,464 (11.83) |

Abbreviations: SD, standard deviation; COPD, chronic obstructive pulmonary disease.
All datasets included 173,371 participants, equivalent to the size of the real UK Biobank training dataset.

**Figure 1.**
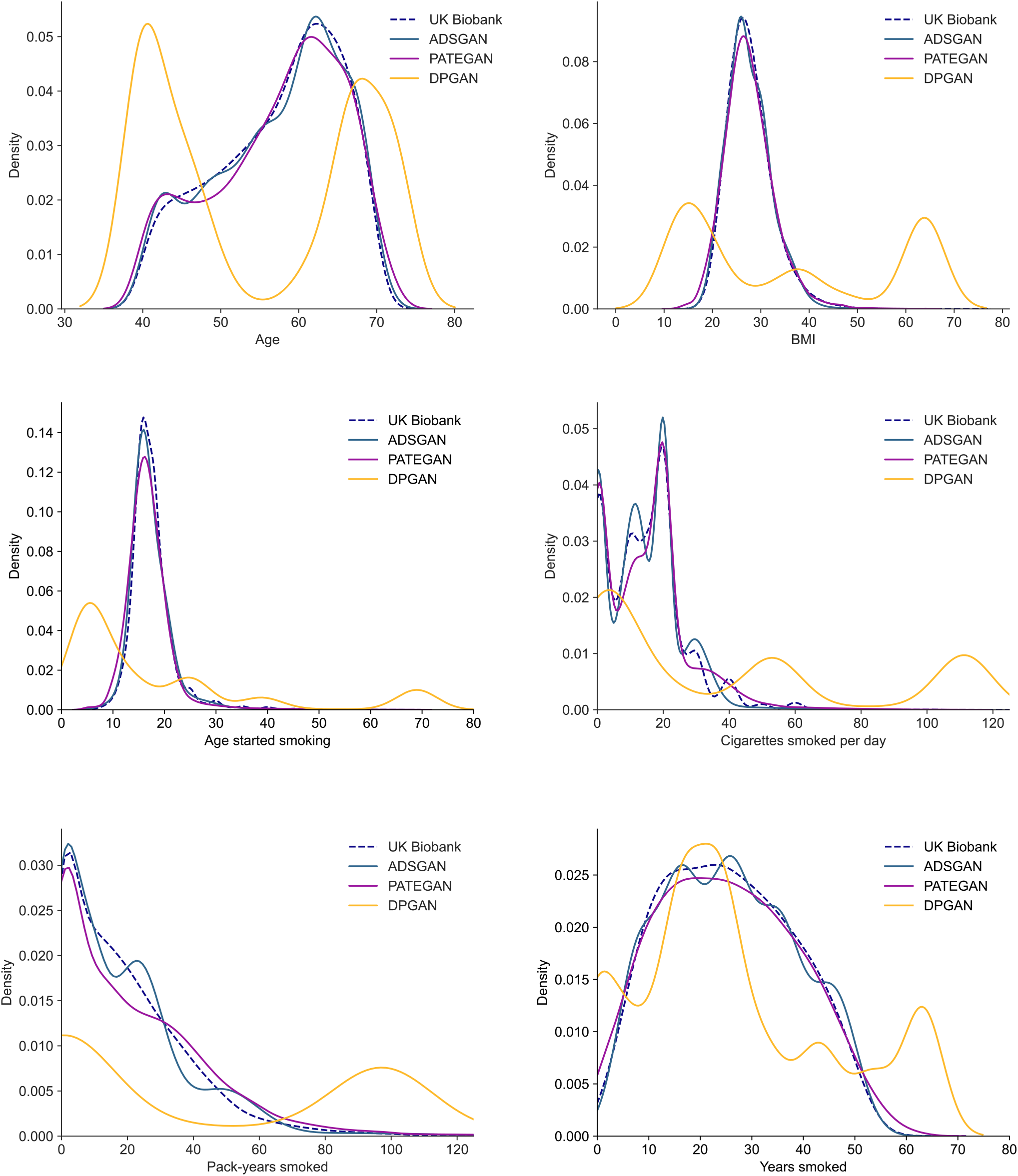
Correspondence between the distribution of continuous variables in the real and synthetic datasets. Synthetic data generated with ADSGAN or PATEGAN maintained the distributions seen in the real training data. DPGAN showed substantial variation from the original and suffered from both mode invention and mode collapse.

#### Principal component analysis

The first step in PCA is to choose the number of principal components, usually by examining the profile of explained variance^40^. As shown in the scree plot in Figure 2a, the variance explained by number of principal components was similar with ADSGAN and DPGAN to the real data. The profile of PATEGAN was different but an important characteristics was shared: most of the variance was explained by the first four components before the curve flattens out. We subsequently fit PCA models using four principal components on the synthetic datasets before evaluating model quality using the log-likelihood in the real test dataset 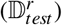. The performance of our PCA model trained on ADSGAN was nearly identical to that trained on the real data 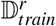 (the “oracle” model), followed closely by PATEGAN (Table 2).

**Figure 2.**
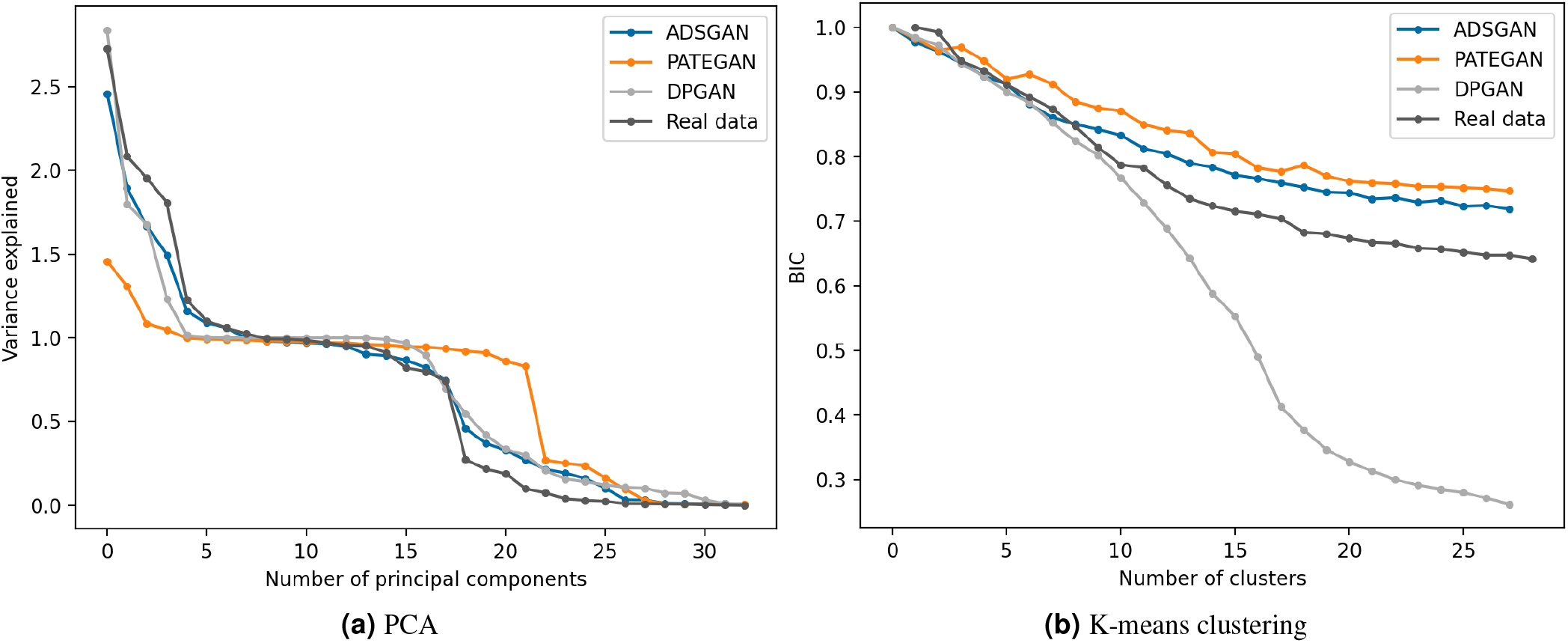
Dimensionality reduction with principal component analysis and K-means clustering in both synthetic and real datasets. (a) shows the variance explained by different numbers of principal components. (b) shows the Bayesian Information Criterion (BIC) of K-means clustering with varying numbers of clusters (indexed at one).

**Table 2.**
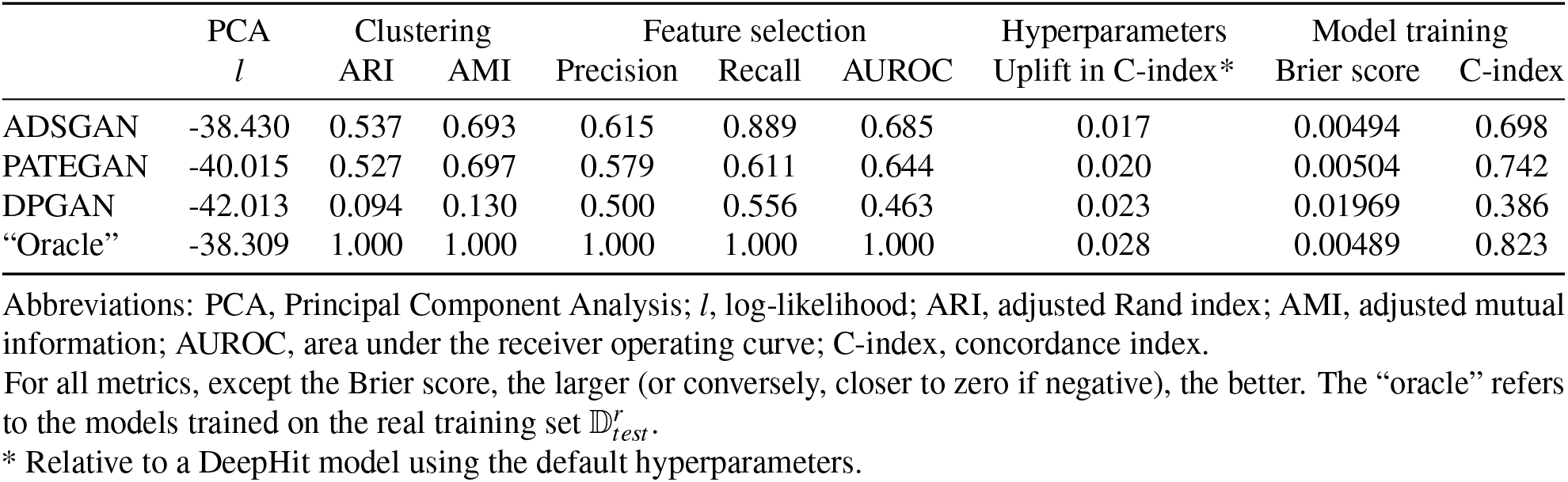
Quantitative evaluation results for different analytical tasks.

#### Clustering with K-means

K-means is a widely used clustering method^39^. Similar to PCA, the number of clusters (*K*) must first be selected. The aim is to find the minimum number of clusters - reducing the dimensions present in the dataset - whilst also reducing the intra-cluster variance. The Bayesian Information Criterion (BIC) is one approach to guide this choice. Figure 2b shows the BIC profile produced with both real and synthetic datasets with respect to the number of clusters. The overall trends in the BIC were similar across the datasets: the curve decreased until reaching its lowest (best) score at 28 clusters. Note, the BIC with DPGAN paralleled that of the real dataset for 10 clusters before significantly diverging. Although the lowest BIC was at 28 clusters in the real and synthetic data generated by ADSGAN and PATEGAN, the rate of decrease in the BIC reduced significantly after 15 clusters across all three datasets. Therefore, an individual with only synthetic data would still be able to use an analysis of BIC to decide on a reasonable number of clusters.

Subsequently, we performed K-means clustering, with *K* = 15, in both the synthetic and real datasets. We used the clusters identified when training a model with the real training dataset, 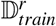, as our comparator “oracle”. We show the agreement between the clusters identified in the real data (the “oracle”) and those derived from the synthetic datasets, evaluated on the test set 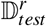 in terms of ARI and AMI in Table 2. Both metrics would be zero if the clusters were randomly assigned and one if the clustering derived from the synthetic data were in perfect agreement with the oracle. Clusters found from synthetic datasets generated with ADSGAN and PATEGAN agreed well with the oracle. However, the synthetic data generated by DPGAN fell short in producing meaningful clusters.

#### Prognostic model development with synthetic data

##### Feature selection

Real world data often contain features that are irrelevant to the prediction task. Here we explored whether feature selection can be reliably performed on synthetic data. The most important features for predicting lung cancer risk in the real data were age, body mass index, smoking duration, pack-years, quit-years, current smoking status, family history of lung cancer and highest qualifications. These features are in keeping with the findings of prior medical literature, with each of the variables included in existing prognostic models for lung cancer^34^.

When performing feature selection with synthetic data, those generated by ADSGAN showed the highest concordance with the real data, keeping all but one of the top ten features: highest qualification (degree). Similarly, feature selection with synthetic data generated by PATEGAN and DPGAN reached similar conclusions, with two discordant features each. The features and associated *p*-values are presented in Appendix Table 2.

To quantify the comparison, Table 2 reports the precision, recall, and AUROC of the true important features when the selection is based on synthetic data. Synthetic data generated by ADSGAN and PATEGAN demonstrated their suitability for feature selection independent of the real dataset, and consistently outperformed those generated by DPGAN.

##### Hyperparameter tuning

Hyperparameter tuning is a complex and important element of prognostic model development and selection. To analyse the reliability of hyperparameter tuning with synthetic data, we trained a neural network-based deep survival model, DeepHit, to predict lung cancer occurrence. There are three key parameters - *α, σ*, and dropout rate - that most significantly impact the performance of DeepHit^45^. By contrast, batch size, hidden dimensionality, learning rate, and patience (for early stopping) are relatively generic deep learning hyperparameters.

The optimal hyperparameters identified when training with real and synthetic datasets were similar across the three key parameters, but with less agreement on the number of hidden dimensions (Table 3). However, this observation is in keeping with prior studies which suggest that the performance of a deep survival model is less sensitive to the number of hidden dimensions^49,50^. We further quantify the usefulness of hyperparameter tuning on synthetic data in Table 2, where we report the improvement in model discrimination (C-index) on the real test dataset 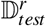. Model tuning in all three synthetic datasets led to a performance gain relative to using the default hyperparameters. These findings suggest that reasonable hyperparameter settings can be identified when using synthetic data that generalize well to real data.

##### Model training

To evaluate the performance of prognostic models trained on synthetic data for real-world deployment without further refitting to real data, we used the train-on-synthetic, test-on-real approach^27^. We developed Cox models using all available features to predict the risk of lung cancer occurrence in each synthetic dataset and in the real training dataset.

Models trained on synthetic data generated by ADSGAN and PATEGAN showed relatively strong discrimination, though less than that achieved when trained on the real data. However, the Brier scores for models trained on ADSGAN-derived synthetic data and real data were equivalent (0.00494 vs 0.00489), with a model trained on PATEGAN-derived synthetic data also performing well. Brier scores quantify the closeness of predicted and observed probabilities and can be decomposed into elements including both calibration and discrimination. This suggests that models trained with ADSGAN and PATEGAN were very well calibrated when tested on real data and able to capture core aspects of the relationship between the variables and the outcome. DPGAN-derived synthetic data were not useful for model development.

Given the trade-off between privacy and utility with synthetic data, a degree of performance drop compared with models trained on real data is unsurprising. However, the strength of the Brier scores suggest that access to synthetic data can inform model development, though further fitting to real data would be necessary to improve their discrimination.

## Discussion

Under real-world assumptions, synthetic data generated with existing privacy-preserving algorithms can be deployed to support all stages of clinical risk prediction modelling. In common with previous analyses, no single generative model was unequivocally best at all tasks. However, in our analyses both ADSGAN and PATEGAN performed consistently well, with limited differences between them across tasks.

We show that synthetic data can be used for exploratory analyses in several ways. First, synthetic datasets preserved the distribution of both continuous and categorical variables from the real dataset. Although seemingly straightforward, substantial insight can be derived from descriptive analyses, with uses ranging from project planning and hypothesis generation, to healthcare operations and logistics. Second, by capturing the underlying characteristics and relationships present within the data, synthetic data can be used to select hyperparameters in unsupervised models, shown here by the number of components in Principal Component Analysis and the Bayesian Information Criterion associated with selecting different numbers of clusters. Indeed, we found that both PCA and K-means clustering performed on purely synthetic data translated well to real datasets.

Building on exploratory analyses, we show the value of synthetic data for feature selection, hyperparameter tuning, and model training under the challenging scenario of right-censored time-to-event analyses using both conventional statistical approaches - Cox models - and deep learning. In these analyses, feature selection based on hypothesis testing using synthetic datasets created with ADSGAN and PATEGAN yielded comparable feature sets to the original dataset. Furthermore, the hyperparameters selected for a deep learning model trained on synthetic data were similar to the real dataset, particularly across those hyperparameters such as model *α, σ*, and dropout rate, that most impact model performance. Finally, Cox models trained on synthetic data had strong Brier scores, approaching that of a model trained on the real dataset, although their discrimination was lower. Given the trade-off between usefulness and privacy, such a drop in performance is expected. Nonetheless, the similarity in Brier scores suggests that synthetic data can be valuable for model development.

Our results have several implications for how synthetic data might be deployed in healthcare settings. Although there are a myriad of different underlying use-cases, how synthetic data could be deployed can largely divided into two approaches: *no-release* or *delayed-release*. Under the most stringent *no-release* situation, the data user has no access to the real data and any analyses they perform on synthetic data will not be validated on the real dataset. Our analyses suggest that synthetic data can still confidently support exploratory data analyses, particularly descriptive analyses, and the planning of further analyses. Nevertheless, the strength of conclusions that can be drawn from prognostic models developed in such a situation will necessarily remain limited. By contrast, multiple use-cases support a *no-release* paradigm where the user has the ability to establish a ground-truth. For example, where the data controller can run code to verify analyses written for synthetic data. In this situation, we show that all aspects of model development could be performed, substantially reducing the risks of data sharing. Furthermore, we also show how synthetic data could support *delayed-release* approaches to data sharing. Through exploratory data analyses and initial model development, a user can ascertain both the suitability of the dataset for the problem they are approaching, and de-risk projects. Subsequently, when the real data become accessible, the user can quickly progress to the application of different modelling approaches.

Synthetic versions of large-scale real-world datasets have been attempted previously for both research-grade primary care data within the UK Clinical Practice Datalink (CPRD)^23^, and the UK National Cancer Registry. However, to date, neither have been able to support use-cases beyond tabulating variable counts, limiting their utility. Consequently, to our knowledge, this is the first work in a clinical context to demonstrate the usability of synthetic data beyond basic descriptive analyses in a complex non-imaging medical dataset.

This work has several limitations. Analyses were performed in one dataset, although the UK Biobank is both large and represents the type of data that is used and shared in a medical context. Further, we curated this dataset and performed imputation prior to synthetic data generation. At present, the generation of high-quality synthetic data in clinical research and development requires such preprocessing. This has advantages in that the data controller will know their data better than any user, but does increase the skillset required by the data controller to generate the synthetic data. Finally, although we show that both ADSGAN and PATEGAN generated high-quality synthetic data, it remains the case that a range of synthetic data generators should be used and evaluated by the data controller before data release. Notably, we found that DPGAN had limited utility. This may reflect the fact that DPGAN is one of the original approaches to integrating differential privacy into synthetic data generation, such that the noise introduced may limit its usefulness at the relatively strong privacy guarantee implied in this analysis.

In summary, synthetic data could be a valuable approach to increasing data sharing, with uses across the whole clinical prognostic modelling pipeline. Whether synthetic data are deployed with or without eventual access to the real data, they can support analyses at all stages from planning to completion, accelerating and de-risking projects, whilst opening new avenues for collaboration and sharing between datasets that have historically remained siloed. Further research to support the deployment of synthetic data in clinical settings should be pursued.

## Data Availability

All data produced in the present study are available upon reasonable request to the authors

https://www.ukbiobank.ac.uk/

## Acknowledgements

This research has been conducted using the UK Biobank Resource under application number 77097. We wish to thank all participants in the UK Biobank and the Biobank coordinating centre.

## Author contributions statement

ZQ, MvdS, and TC conceived the study. ZQ and TC performed the analyses and interpreted the results. Specifically, ZQ performed exploratory data analyses and prognostic model development and supported with the generation of synthetic data; TC performed data extraction, imputation, and preprocessing before generating the synthetic datasets and running descriptive analyses. ZQ and TC drafted the manuscript; all authors were involved in manuscript revision. ZQ, TC, NN, and MvdS had full access to the data in the study and can take responsibility for the integrity of the data and the accuracy of the data analysis. TC, NN, and MvdS obtained funding for this project; TC led ethics appproval.

## Ethics

Ethical approval was granted for this project by the HRA and Health and Care Research Wales (HCRW) approval board (reference: 21/LO/0779).

