## Appendix for "Synthetic data for privacy-preserving clinical risk prediction"

### Appendix Tables

1 Candidate variables included in the synthetic data . . . . . 2
2 Feature selection with real and synthetic datasets. . . . . . 3
3 Hyperparameter configurations of DeepHit identified from real and synthetic datasets. . . . . 3

**Appendix Table 1.** Candidate variables included in the synthetic data

| Candidate variables |
| --- |
| Age |
| Sex |
| Body mass index |
| Ethnicity |
| Highest qualification |
| Age started smoking |
| Age stopped smoking |
| Smoking duration |
| Years since stopped smoking |
| Pack-years smoked |
| Smoking status |
| Asbestos exposure |
| Personal history of asbestosis |
| Personal history of pneumonia |
| Personal history of COPD |
| Personal history of Emphysema |
| Personal history of chronic bronchitis |
| Personal history of asthma |
| Personal history of eczema, allergic rhinitis, or hayfever |
| Personal history of cancer |
| Number of previous cancers |
| Family history of lung cancer (father) |
| Family history of lung cancer (mother) |
| Family history of lung cancer (siblings) |

**Appendix Table 2.** Feature selection with real and synthetic datasets.

| Variable | Real | ADSGAN | PATEGAN | DPGAN |
|---|---|---|---|---|
| Smoking duration (years) | 1.3E-58 | 7.5E-03 | 9.4E-271 | 1.2E-32 |
| Age | 2.3E-50 | 1.7E-15 | 5.3E-159 | 6.0E-178 |
| Pack-years | 5.4E-29 | 5.1E-20 | 6.0E-09 | 0.0E+00 |
| Years since stopped smoking | 1.8E-12 | 1.2E-36 | 4.2E-09 | 0.0E+00 |
| Current smoking status | 4.6E-11 | 5.9E-13 | 8.4E-05 | 1.3E-83 |
| Family history of lung cancer (father) | 1.4E-07 | 9.2E-05 | 1.6E-03 | - |
| Family history of lung cancer (siblings) | 5.0E-07 | 8.8E-09 | - | - |
| Highest qualifications - degree | 3.0E-05 | - | 3.2E-08 | 1.1E-110 |
| Highest qualification - other | 2.0E-10 | 1.9E-04 | - | 3.1E-03 |
| Body mass index | 3.7E-05 | 3.3E-66 | 1.5E-98 | 1.1E-57 |

P values of the top ten features calculated on different data sets. "-" indicates p value > 0.05.

**Appendix Table 3.** Hyperparameter configurations of DeepHit identified from real and synthetic datasets.

| | $\alpha$ | $\sigma$ | Dropout | Batch size | Hidden dim | Learning rate | Patience |
|---|---|---|---|---|---|---|---|
| Real | 0.358 | 0.358 | 0.143 | 200 | 10 | 0.001 | 13 |
| ADSGAN | 0.323 | 0.323 | 0.129 | 500 | 100 | 0.0001 | 29 |
| PATEGAN | 0.264 | 0.264 | 0.106 | 500 | 70 | 0.0001 | 34 |
| DPGAN | 0.463 | 0.463 | 0.185 | 500 | 90 | 0.0001 | 11 |
